# Association between Physical Function and Incidence of Atrial Fibrillation: The Atherosclerosis Risk in Communities (ARIC) Study

**DOI:** 10.64898/2026.04.13.26350644

**Authors:** Byung Joon Pae, Linzi Li, Kathryn Wood, Elsayed Z. Soliman, Lin Yee Chen, Faye L. Norby, B. Gwen Windham, Alvaro Alonso

## Abstract

**Background:** Poor physical function has been associated with higher cardiovascular disease (CVD) risk. However, the association between physical function and atrial fibrillation (AF) remains understudied. The comprehensive investigation of the association between physical function and incident AF risk could highlight a novel target for AF prevention.

**Methods:** A total of 4,803 participants without diagnosed AF from the Atherosclerosis Risk in Communities (ARIC) Study cohort with physical function assessed in 2011-2013 were studied. Physical function was measured using Short Physical Performance Battery (SPPB), 4-meter walk time, and grip strength. Hospital discharge codes and death certificates were used to ascertain incident AF through 2022, and through 2020 for participants from Jackson. Cox regression was used to assess the association between physical function and incident AF risk, adjusting for multiple covariates. Z-score transformations were performed to identify the physical function measure most strongly associated with incident AF risk, and SPPB component analysis was performed to identify the most influential SPPB component.

**Results:** Mean age of the study participants was 75.1 ± 5.0 years, with 41.2% being male participants and 22.2% being black participants. During a median follow-up of 9.2 years, there were 809 incident AF events. SPPB (HR: 0.93, 95% CI: 0.90-0.96, per 1-point increase) and grip strength (HR: 0.87, 95% CI: 0.78-0.96, per 10kg increase) were inversely associated with incident AF risk, while 4-meter walk time (HR: 1.08, 95% CI: 1.03-1.13, per 1-second increase) was positively associated with incident AF risk. SPPB had the strongest association with incident AF risk. Within SPPB, only the chair stand component was significantly associated with incident AF risk.

**Conclusions:** The findings suggest that better physical function is associated with reduced incident AF risk, with higher SPPB having the strongest association. Given the modifiable nature of physical function, these findings highlight a potential novel target for AF prevention in aging populations.

**What is Known:**

- Physical function has been associated with cardiovascular diseases, however, the relationship between physical function and incident atrial fibrillation (AF) remains understudied.

**What the Study Adds:**

- This study found that better Short Physical Performance Battery (SPPB), 4-meter walk time, and grip strength were all independently associated with reduced risk of incident AF.
- In this study, higher SPPB was most strongly associated with reduced risk of incident AF, implying the importance of multi-domain measures of physical function.
- This study found that within SPPB, higher chair stand component score was the only component significantly associated with reduced risk of incident AF, highlighting the critical role of muscle strength in the association between physical function and risk of incident AF.
- The results suggest that physical function may be a novel modifiable target for AF prevention.

## Introduction

Atrial Fibrillation (AF) is the most common clinically significant arrhythmia and continues to grow globally.^1, 2^ The Global Burden of Disease study estimated the global AF prevalence at 46.3 million in 2016 and 59 million in 2019.^3, 4^ Evidence also demonstrates a 4-fold increase in age-standardized AF prevalence over the last 50 years.^5^ Considering that many individuals are not diagnosed with AF until symptoms manifest, these figures likely underestimate the true prevalence of AF.^1, 6^ AF is associated with increased mortality and augmented risk of developing deleterious conditions such as myocardial infarction, stroke, heart failure, and dementia.^1, 2, 7, 8, 9, 10^ Thus, it is imperative to identify interventions for the prevention of AF.

In recent years, physical function has emerged as an independent predictor of cardiovascular disease (CVD).^11, 12, 13, 14, 15, 16, 17, 18, 19, 20, 21^ Such physical function parameters include, but are not limited to muscle strength, gait speed, and the Short Physical Performance Battery (SPPB). Despite the accumulation of studies suggesting the association between physical function and CVD, the association between physical function and AF specifically is understudied. A previous study reported an association between handgrip strength, a measure of physical function, and AF risk, but it did not consider other physical function measurements.^22^ The comprehensive investigation of the association between several physical function parameters and risk of incident AF is needed, as it could potentially identify modifiable targets for AF prevention and novel mechanisms related to AF risk.

This study aims to assess the association between various measures of physical function and risk of incident AF and identify the measure of physical function that is most strongly associated with risk of incident AF. We hypothesize that better physical function will be associated with reduced risk of incident AF, and higher SPPB will be most strongly associated with reduced risk of incident AF, as it is a multi-domain measure of physical function.

## Methods

### Study Participants

Data from the Atherosclerosis Risk in Communities (ARIC) Study were used. Briefly, the ARIC Study is an ongoing community-based prospective cohort study that was designed to identify risk factors for various CVDs.^23^ The study enrolled 15,792 participants aged 45 to 64 in 1987-1989 (Visit 1) from 4 US communities: Washington County, MD; Forsyth County, NC; Jackson, MS; and selected suburbs of Minneapolis, MN. Participants received periodic in-person follow-up examinations and were monitored for cardiovascular outcomes. The Visit 5 examinations, conducted between 2011 and 2013, comprised 6,538 participants and was used as the baseline for this study. Due to insufficient sample sizes, (1) non-White participants from the Minneapolis and Washington County field centers and (2) non-White and non-Black participants from the Forsyth County field center were excluded. Jackson field center was restricted to Black participants; therefore, no race-based exclusions were applied. Participants with a history of AF at or prior to baseline, missing physical function data, and missing covariate data were further excluded. History of AF at or prior to baseline was determined by electrocardiograms and hospital discharge codes (ICD-9-CM: 427.3x). 4,803 participants were included in the analysis (Figure 1). All participants provided informed consent, and the research protocol was approved by the Institutional Review Boards of all participating institutions.

**Figure 1.**
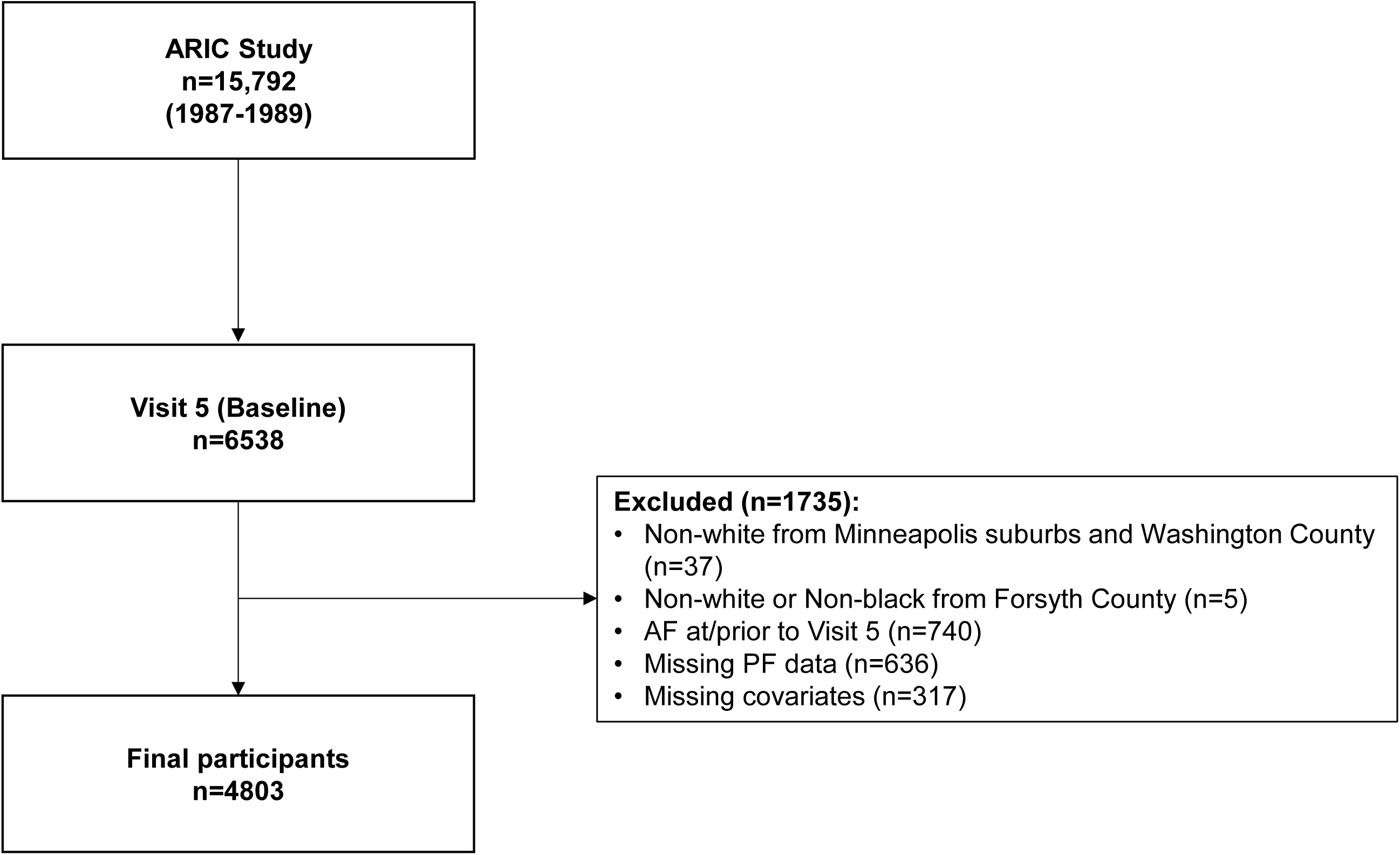
Flowchart of the selection of study participants. ARIC, atherosclerosis risk in communities; AF, atrial fibrillation; PF, physical function.

## Physical Function

### Short Physical Performance Battery (SPPB)

Lower extremity physical function was assessed using the Short Physical Performance Battery (SPPB).^24^ Briefly, the SPPB consists of 3 components: 4-meter walk; standing balance; and chair stand. Each component is scored from 0 to 4. The SPPB total score is the sum of the 3 component scores and ranges from 0 to 12, with higher scores reflecting greater physical function.

Regarding the 4-meter walk component, participants were instructed to walk 4-meters at their usual pace twice, and the faster result was recorded. The 4-meter walk component was scored based on the time to complete 4-meters: 0 (unable to walk 4 m); 1 (>8.70 seconds); 2 (≥6.21 to ≤8.70 seconds); 3 (≥4.82 to <6.21 seconds); and 4 (<4.82 seconds).

The standing balance component required the participants to maintain balance for 10 seconds while being in the side-by-side, semi-tandem, and tandem position. The assessment started with the semi-tandem position. If participants were unable to perform the semi-tandem position, the side-by-side position was performed. For the semi-tandem and side-by-side positions, participants received a score of 1 for completion; otherwise, a score of 0 was given. The tandem position was scored based on the time able to maintain the position: 0 (≤3 seconds); 1 (>3 to <10 seconds); and 2 (≥10 seconds). The standing balance component score is the sum of all three positions, ranging from 0 to 4.

For the chair stand component, participants were asked to fold their arms across the chest and perform 5 chair stands using a chair. The chair stand component score was based on the time to complete 5 chair stands: 0 (unable to finish in 60 seconds); 1 (≥16.7 to <60 seconds); 2 (≥13.7 to <16.7 seconds); 3 (≥11.2 to <13.7 seconds); and 4 (<11.2 seconds).

### 4-Meter Walk Time

As the 4-meter walk component score from the SPPB was scored from 0 to 4, the unprocessed time to complete 4-meters was analyzed separately as a continuous variable.

### Grip Strength

Upper extremity physical function was assessed using grip strength. Grip strength (kg) was measured twice for each participant using the preferred hand. The better result of the 2 trials was used.^25^

### Incidence of Atrial Fibrillation

The outcome of this study was incident AF, defined as any AF cases that occur following the Visit 5 examination (2011-2013) and prior to December 31, 2022; however, incident AF events for participants from Jackson were ascertained through December 31, 2020. AF was defined from hospital discharge codes (ICD-9-CM: 427.3x; ICD-10-CM: I48.x) not occurring in the context of open cardiac surgery and from death certificates with AF as an underlying or contributing cause of death (ICD-10: I48).^26^

### Covariates

Covariates included age, sex, race, center, education, body mass index (BMI), alcohol drinking status, smoking status, leisure time sport-related physical activity, leisure time physical activity excluding sport, total cholesterol, low-density lipoprotein cholesterol (LDL), high-density lipoprotein cholesterol (HDL), triglyceride, hypertension, diabetes, and history of cardiovascular diseases (stroke, heart failure, coronary heart disease). Self-reported information on sex, race, center, and education were collected at Visit 1. Education was stratified into 6 levels: grade school of 0 years; high school (no degree); high school (graduate); vocational school; college; and graduate/professional school. The college category did not require a completed degree. Other covariates were measured at Visit 5. BMI was defined as the weight (kg) divided by the square of height (m^2^). Drinking and smoking were self-reported and categorized into current, former, never, and unknown. Leisure time sport-related physical activity and leisure time physical activity excluding sport were self-reported and measured by an interviewer using the Baecke Questionnaire.^27^ Both physical activity level scores range from 1 to 5, with higher scores indicating greater physical activity levels. Total cholesterol, LDL, HDL, and triglycerides were assessed using standardized procedures in blood samples collected at the visit. Hypertension was defined as systolic blood pressure ≥ 140 mmHg, diastolic blood pressure ≥ 90 mmHg, or use of medication for high blood pressure. Diabetes was defined as fasting glucose level ≥ 126 mg/dL, non-fasting glucose level ≥ 200 mg/dL, use of medication for diabetes, or self-reported diagnosis of diabetes by a physician. History of stroke, HF, and CHD were defined based on the ARIC criteria.^28, 29, 30^

### Statistical Analysis

Cumulative incidence of AF was estimated using the Kaplan-Meier method and compared between high and low SPPB groups, which was determined by the SPPB total score (< median vs. ≥ median). Baseline characteristics were compared between the aforementioned two groups. Continuous variables were reported as mean (SD) and compared using Student’s t test or Mann-Whitney U test, depending on normality, and categorical variables were reported as count (%) and compared using the χ^2^ test. Cox regression was utilized for assessing the association between physical function and risk of incident AF, with follow-up time defined as time from baseline to AF incidence, censoring due to death or loss to follow-up, or 12/31/2022, whichever is earlier.

For participants from Jackson, follow-up time was defined as time from baseline to AF incidence, censoring due to death or loss to follow-up, or 12/31/2020, whichever is earlier. Model 1 was a crude model. Model 2 adjusted for age, sex, race, and center, accounting for the study’s race and center specific recruitment structure. Model 3 further adjusted for education, BMI, alcohol drinking status, smoking status, leisure time sport-related physical activity, leisure time physical activity excluding sport, total cholesterol, LDL, HDL, triglyceride, hypertension, diabetes, and history of cardiovascular diseases (stroke, heart failure, coronary heart disease).

SPPB was analyzed as both continuous and categorical variables.^11^ The categorical SPPB was stratified into low (0-6), intermediate (7-9), and high (10-12) physical function.^11, 31^ Grip strength was divided by 10 to represent per 10 kg increase. 4-meter walk time in seconds was analyzed in its original form. All measures of physical function were tested separately. As previous studies have reported sex and race differences in AF, additional analyses stratified by sex and race were performed.^32, 33^

Restricted cubic splines were used to assess nonlinearity and dose-response relationship between physical function and risk of incident AF.^34^ The number of knots was determined by lowest Akaike Information Criterion (AIC). For determining the measure of physical function most strongly associated with risk of incident AF, z-score transformations were performed, and all transformed measures of physical function were tested together in the same model. The SPPB component analysis was performed to assess which component within SPPB is most strongly associated with risk of incident AF and tested all 3 components (4-meter walk, standing balance, and chair stand) together in the same model. Two-sided p-value < 0.05 was considered statistically significant. All statistical analyses were performed using SAS software (Version 9.4; SAS Institute, Cary, NC, US).

## Results

### General Characteristics

Table 1 displays the general characteristics at baseline. Of the 4,803 participants included for analysis, the mean age was 75.1 ± 5.0 years with 41.2% being male participants and 22.2% being black participants. The median SPPB was 10. High (≥10) SPPB included 2,852 participants (age, 74.2 ± 4.5 years; 45.7% males; 16.3% blacks) while 1,951 participants were considered as low SPPB (age, 76.6 ± 5.3 years; 34.6% males; 30.8% blacks). The low (<10) SPPB group had a greater proportion of lower education, current smokers, hypertension, diabetes, stroke history, heart failure history, and coronary heart disease history compared to the high SPPB group.

**Table 1.**
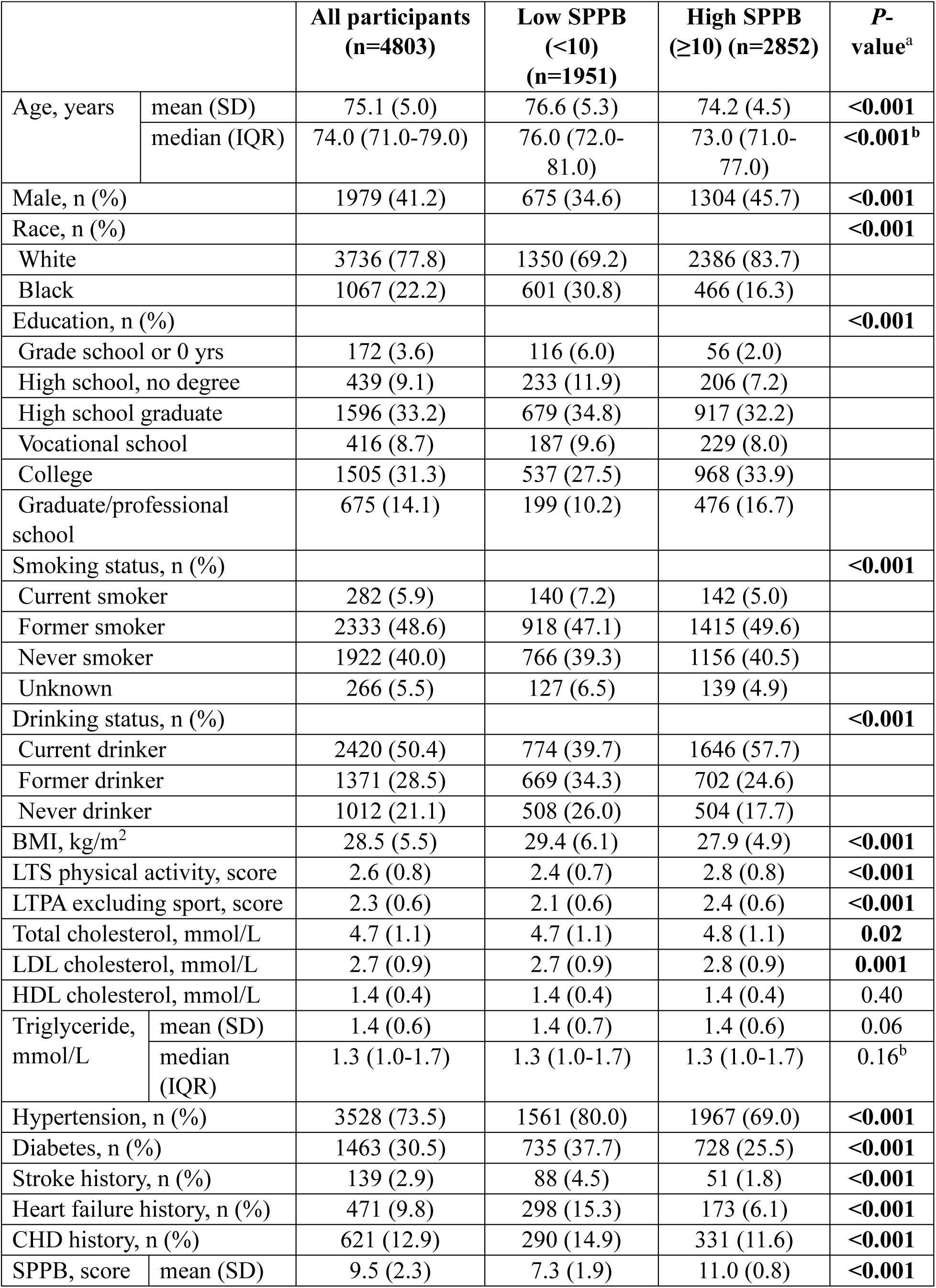
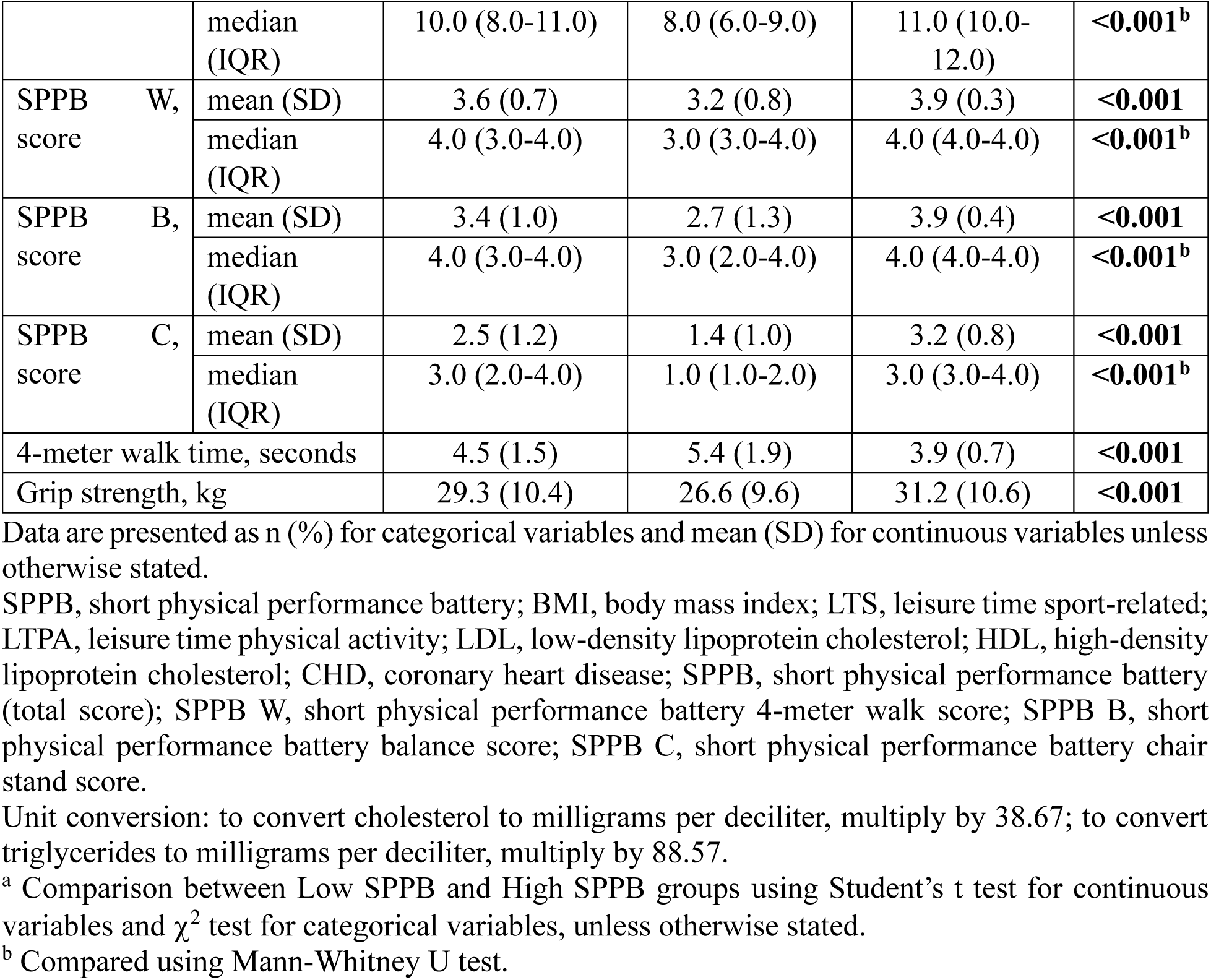
Baseline Characteristics Overall and by SPPB Levels, ARIC Cohort, 2011-2013.

|  |  | All participants<br>(n=4803) | Low SPPB<br>(<10)<br>(n=1951) | High SPPB<br>(≥10) (n=2852) | P-value<sup>a</sup> |
| --- | --- | --- | --- | --- | --- |
| Age, years | mean (SD) | 75.1 (5.0) | 76.6 (5.3) | 74.2 (4.5) | <0.001 |
|  | median (IQR) | 74.0 (71.0-79.0) | 76.0 (72.0-81.0) | 73.0 (71.0-77.0) | <0.001<sup>b</sup> |
| Male, n (%) |  | 1979 (41.2) | 675 (34.6) | 1304 (45.7) | <0.001 |
| Race, n (%) |  |  |  |  | <0.001 |
| White |  | 3736 (77.8) | 1350 (69.2) | 2386 (83.7) |  |
| Black |  | 1067 (22.2) | 601 (30.8) | 466 (16.3) |  |
| Education, n (%) |  |  |  |  | <0.001 |
| Grade school or 0 yrs |  | 172 (3.6) | 116 (6.0) | 56 (2.0) |  |
| High school, no degree |  | 439 (9.1) | 233 (11.9) | 206 (7.2) |  |
| High school graduate |  | 1596 (33.2) | 679 (34.8) | 917 (32.2) |  |
| Vocational school |  | 416 (8.7) | 187 (9.6) | 229 (8.0) |  |
| College |  | 1505 (31.3) | 537 (27.5) | 968 (33.9) |  |
| Graduate/professional school |  | 675 (14.1) | 199 (10.2) | 476 (16.7) |  |
| Smoking status, n (%) |  |  |  |  | <0.001 |
| Current smoker |  | 282 (5.9) | 140 (7.2) | 142 (5.0) |  |
| Former smoker |  | 2333 (48.6) | 918 (47.1) | 1415 (49.6) |  |
| Never smoker |  | 1922 (40.0) | 766 (39.3) | 1156 (40.5) |  |
| Unknown |  | 266 (5.5) | 127 (6.5) | 139 (4.9) |  |
| Drinking status, n (%) |  |  |  |  | <0.001 |
| Current drinker |  | 2420 (50.4) | 774 (39.7) | 1646 (57.7) |  |
| Former drinker |  | 1371 (28.5) | 669 (34.3) | 702 (24.6) |  |
| Never drinker |  | 1012 (21.1) | 508 (26.0) | 504 (17.7) |  |
| BMI, kg/m<sup>2</sup> |  | 28.5 (5.5) | 29.4 (6.1) | 27.9 (4.9) | <0.001 |
| LTS physical activity, score |  | 2.6 (0.8) | 2.4 (0.7) | 2.8 (0.8) | <0.001 |
| LTPA excluding sport, score |  | 2.3 (0.6) | 2.1 (0.6) | 2.4 (0.6) | <0.001 |
| Total cholesterol, mmol/L |  | 4.7 (1.1) | 4.7 (1.1) | 4.8 (1.1) | 0.02 |
| LDL cholesterol, mmol/L |  | 2.7 (0.9) | 2.7 (0.9) | 2.8 (0.9) | 0.001 |
| HDL cholesterol, mmol/L |  | 1.4 (0.4) | 1.4 (0.4) | 1.4 (0.4) | 0.40 |
| Triglyceride, mmol/L | mean (SD) | 1.4 (0.6) | 1.4 (0.7) | 1.4 (0.6) | 0.06 |
|  | median (IQR) | 1.3 (1.0-1.7) | 1.3 (1.0-1.7) | 1.3 (1.0-1.7) | 0.16<sup>b</sup> |
| Hypertension, n (%) |  | 3528 (73.5) | 1561 (80.0) | 1967 (69.0) | <0.001 |
| Diabetes, n (%) |  | 1463 (30.5) | 735 (37.7) | 728 (25.5) | <0.001 |
| Stroke history, n (%) |  | 139 (2.9) | 88 (4.5) | 51 (1.8) | <0.001 |
| Heart failure history, n (%) |  | 471 (9.8) | 298 (15.3) | 173 (6.1) | <0.001 |
| CHD history, n (%) |  | 621 (12.9) | 290 (14.9) | 331 (11.6) | <0.001 |
| SPPB, score | mean (SD) | 9.5 (2.3) | 7.3 (1.9) | 11.0 (0.8) | <0.001 |
|  | median (IQR) | 10.0 (8.0-11.0) | 8.0 (6.0-9.0) | 11.0 (10.0-12.0) | **<0.001<sup>b</sup>** |
| SPPB W, score | mean (SD) | 3.6 (0.7) | 3.2 (0.8) | 3.9 (0.3) | **<0.001** |
|  | median (IQR) | 4.0 (3.0-4.0) | 3.0 (3.0-4.0) | 4.0 (4.0-4.0) | **<0.001<sup>b</sup>** |
| SPPB B, score | mean (SD) | 3.4 (1.0) | 2.7 (1.3) | 3.9 (0.4) | **<0.001** |
|  | median (IQR) | 4.0 (3.0-4.0) | 3.0 (2.0-4.0) | 4.0 (4.0-4.0) | **<0.001<sup>b</sup>** |
| SPPB C, score | mean (SD) | 2.5 (1.2) | 1.4 (1.0) | 3.2 (0.8) | **<0.001** |
|  | median (IQR) | 3.0 (2.0-4.0) | 1.0 (1.0-2.0) | 3.0 (3.0-4.0) | **<0.001<sup>b</sup>** |
| 4-meter walk time, seconds |  | 4.5 (1.5) | 5.4 (1.9) | 3.9 (0.7) | **<0.001** |
| Grip strength, kg |  | 29.3 (10.4) | 26.6 (9.6) | 31.2 (10.6) | **<0.001** |

Data are presented as n (%) for categorical variables and mean (SD) for continuous variables unless otherwise stated.
SPPB, short physical performance battery; BMI, body mass index; LTS, leisure time sport-related; LTPA, leisure time physical activity; LDL, low-density lipoprotein cholesterol; HDL, high-density lipoprotein cholesterol; CHD, coronary heart disease; SPPB, short physical performance battery (total score); SPPB W, short physical performance battery 4-meter walk score; SPPB B, short physical performance battery balance score; SPPB C, short physical performance battery chair stand score.
Unit conversion: to convert cholesterol to milligrams per deciliter, multiply by 38.67; to convert triglycerides to milligrams per deciliter, multiply by 88.57.
<sup>a</sup> Comparison between Low SPPB and High SPPB groups using Student's t test for continuous variables and $\chi^2$ test for categorical variables, unless otherwise stated.
<sup>b</sup> Compared using Mann-Whitney U test.

Furthermore, the low SPPB group had a greater BMI and performed less physical activity.

### Incidence of AF

During a median follow-up of 9.2 years, there were 809 incident AF events. The proportion of participants with incident AF events was higher in the low SPPB group (19.7%) compared to the high SPPB group (14.9%) (Figure 2). The overall incidence rate of AF was 21.6 cases per 1,000 person-years. A higher incidence rate of AF was observed in the low SPPB group (27.3 cases per 1,000 person-years) compared to the high SPPB group (18.1 cases per 1,000 person-years).

**Figure 2.**
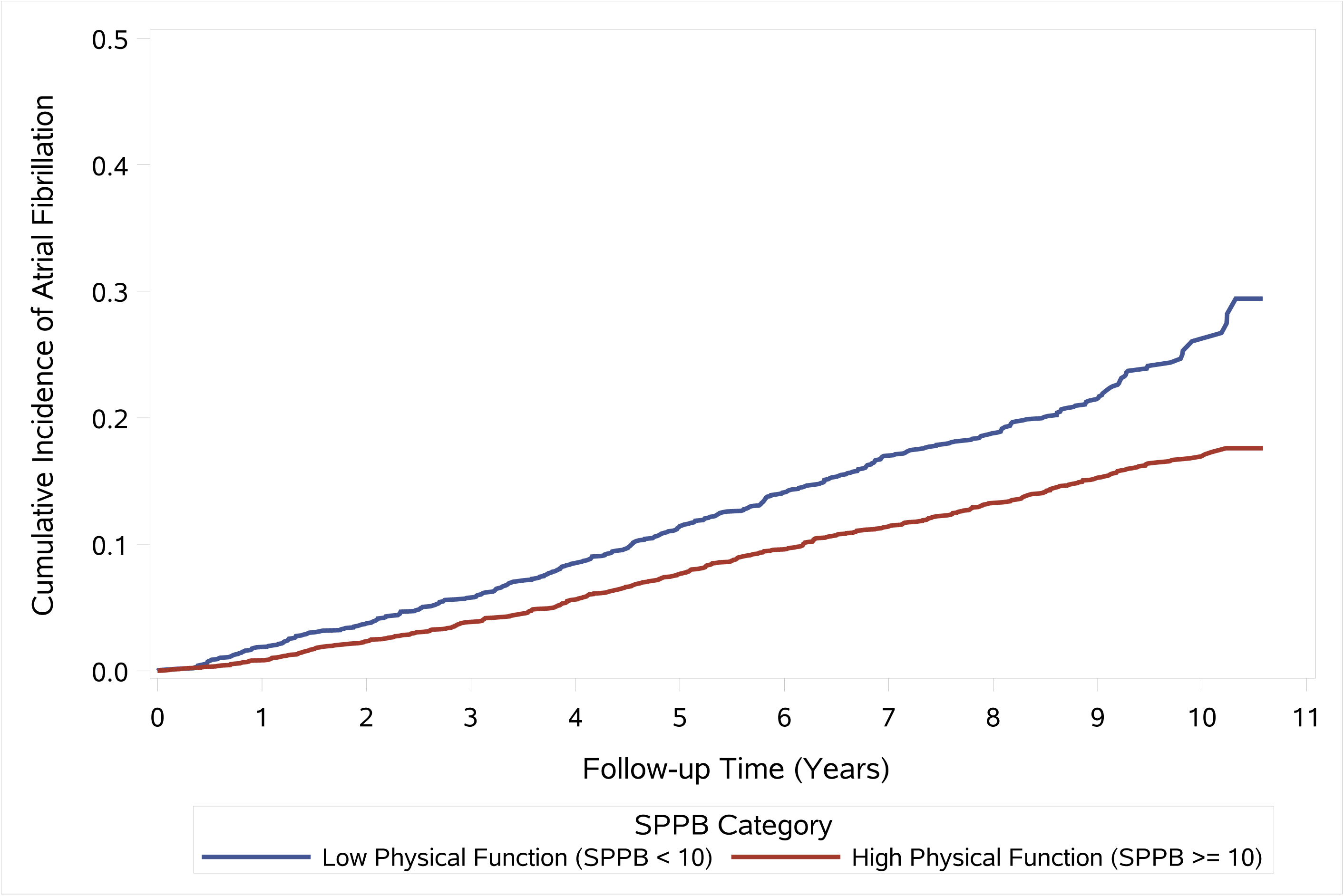
Cumulative incidence of atrial fibrillation by low (SPPB < 10) and high (SPPB >= 10) physical function, estimated using the Kaplan-Meier method. SPPB, short physical performance battery.

### Association between Physical Function and Incident AF

In fully adjusted models (model 3), SPPB (HR: 0.93, 95% CI: 0.90-0.96, p-value: <0.001, 7% risk reduction per 1-point increase), 4-meter walk time (HR: 1.08, 95% CI: 1.03-1.13, p-value: 0.001, 8% increased risk per 1-second increase), and grip strength (HR: 0.87, 95% CI: 0.78-0.96, p-value: 0.006, 13% risk reduction per 10 kg increase) were significantly associated with risk of incident AF (Table 2). Such associations were of similar magnitude in models 1 and 2. The association between SPPB and risk of incident AF was also significant when SPPB was treated as a categorical variable. Compared to low physical function, intermediate (HR: 0.69, 95% CI: 0.55-0.87, p-value: 0.001) and high physical function (HR: 0.61, 95% CI: 0.48-0.77, p-value: <0.001) were significantly associated with reduced risk of incident AF after adjusting for covariates (Table 3). When testing all three measures of physical function together following z-score transformations, SPPB was the only measurement significantly associated with risk of incident AF (HR: 0.86, 95% CI: 0.78-0.95, p-value: 0.002) (Table 4), with no significant associations for 4-meter walk time (HR: 1.02, 95% CI: 0.93-1.12, p-value: 0.64) and grip strength (HR: 0.90, 95% CI: 0.81-1.00, p-value: 0.06). Within SPPB, higher chair stand component score was the only component that was significantly associated with reduced risk of incident AF (Table 5). Compared to a chair stand component score of 0, a score of 4 was associated with a 50% reduced risk of incident AF after adjusting for all covariates (HR: 0.50, 95% CI: 0.36-0.70, p-value: <0.001).

**Table 2.** Association between Measures of Physical Function and Risk of Incident Atrial Fibrillation.

|  | Model 1<sup>a</sup> |  |  | Model 2<sup>b</sup> |  |  | Model 3<sup>c</sup> |  |  |
| --- | --- | --- | --- | --- | --- | --- | --- | --- | --- |
|  | HR | 95% CI | *P*-value | HR | 95% CI | *P*-value | HR | 95% CI | *P*-value |
| SPPB, score | 0.89 | 0.87-0.92 | **<0.001** | 0.89 | 0.86-0.92 | **<0.001** | 0.93 | 0.90-0.96 | **<0.001** |
| 4-meter walk time, seconds | 1.12 | 1.08-1.16 | **<0.001** | 1.13 | 1.09-1.17 | **<0.001** | 1.08 | 1.03-1.13 | **0.001** |
| Grip strength, 10 kg | 0.93 | 0.87-1.00 | **0.04** | 0.81 | 0.74-0.90 | **<0.001** | 0.87 | 0.78-0.96 | **0.006** |

Each measure of physical function was tested separately.
HR, hazard ratio; CI, confidence interval; SPPB, short physical performance battery (total score).
<sup>a</sup> Crude model.
<sup>b</sup> Adjusted for age, sex, race, and center.
<sup>c</sup> Adjusted for age, sex, race, center, education, BMI, drinking status, smoking status, leisure time sport-related physical activity, leisure time physical activity excluding sport, total cholesterol, LDL cholesterol, HDL cholesterol, triglyceride, hypertension, diabetes, history of stroke, history of heart failure, and history of coronary heart disease.

**Table 3.** Association between Short Physical Performance Battery Total Score Categories of Physical Function and Risk of Incident Atrial Fibrillation.

|  | Low (0-6) |  |  | Intermediate (7-9) |  |  | High (10-12) |  |  |
| --- | --- | --- | --- | --- | --- | --- | --- | --- | --- |
|  | HR | 95% CI | P-value | HR | 95% CI | P-value | HR | 95% CI | P-value |
| Model 1<sup>a</sup> | Reference |  |  | 0.63 | 0.51-0.78 | <0.001 | 0.46 | 0.38-0.56 | <0.001 |
| Model 2<sup>b</sup> | Reference |  |  | 0.61 | 0.49-0.76 | <0.001 | 0.46 | 0.37-0.58 | <0.001 |
| Model 3<sup>c</sup> | Reference |  |  | 0.69 | 0.55-0.87 | 0.001 | 0.61 | 0.48-0.77 | <0.001 |

HR, hazard ratio; CI, confidence interval.
<sup>a</sup> Crude model.
<sup>b</sup> Adjusted for age, sex, race, and center.
<sup>c</sup> Adjusted for age, sex, race, center, education, BMI, drinking status, smoking status, leisure time sport-related physical activity, leisure time physical activity excluding sport, total cholesterol, LDL cholesterol, HDL cholesterol, triglyceride, hypertension, diabetes, history of stroke, history of heart failure, and history of coronary heart disease.

**Table 4.** Comparison of Association between Z-Score Transformed Measures of Physical Function and Risk of Incident Atrial Fibrillation.

|  | Model 1<sup>a</sup> |  |  | Model 2<sup>b</sup> |  |  | Model 3<sup>c</sup> |  |  |
| --- | --- | --- | --- | --- | --- | --- | --- | --- | --- |
|  | HR | 95% CI | *P*-value | HR | 95% CI | *P*-value | HR | 95% CI | *P*-value |
| SPPB, score | 0.78 | 0.71-0.85 | **<0.001** | 0.80 | 0.73-0.88 | **<0.001** | 0.86 | 0.78-0.95 | **0.002** |
| 4-meter walk time, seconds | 1.02 | 0.94-1.11 | 0.63 | 1.05 | 0.96-1.14 | 0.28 | 1.02 | 0.93-1.12 | 0.64 |
| Grip strength, 10 kg | 0.99 | 0.92-1.07 | 0.83 | 0.88 | 0.79-0.98 | **0.02** | 0.90 | 0.81-1.00 | 0.06 |

All 3 measures of physical function were tested together.
HR, hazard ratio; CI, confidence interval; SPPB, short physical performance battery (total score).
<sup>a</sup> Crude model.
<sup>b</sup> Adjusted for age, sex, race, and center.
<sup>c</sup> Adjusted for age, sex, race, center, education, BMI, drinking status, smoking status, leisure time sport-related physical activity, leisure time physical activity excluding sport, total cholesterol, LDL cholesterol, HDL cholesterol, triglyceride, hypertension, diabetes, history of stroke, history of heart failure, and history of coronary heart disease.

**Table 5.**
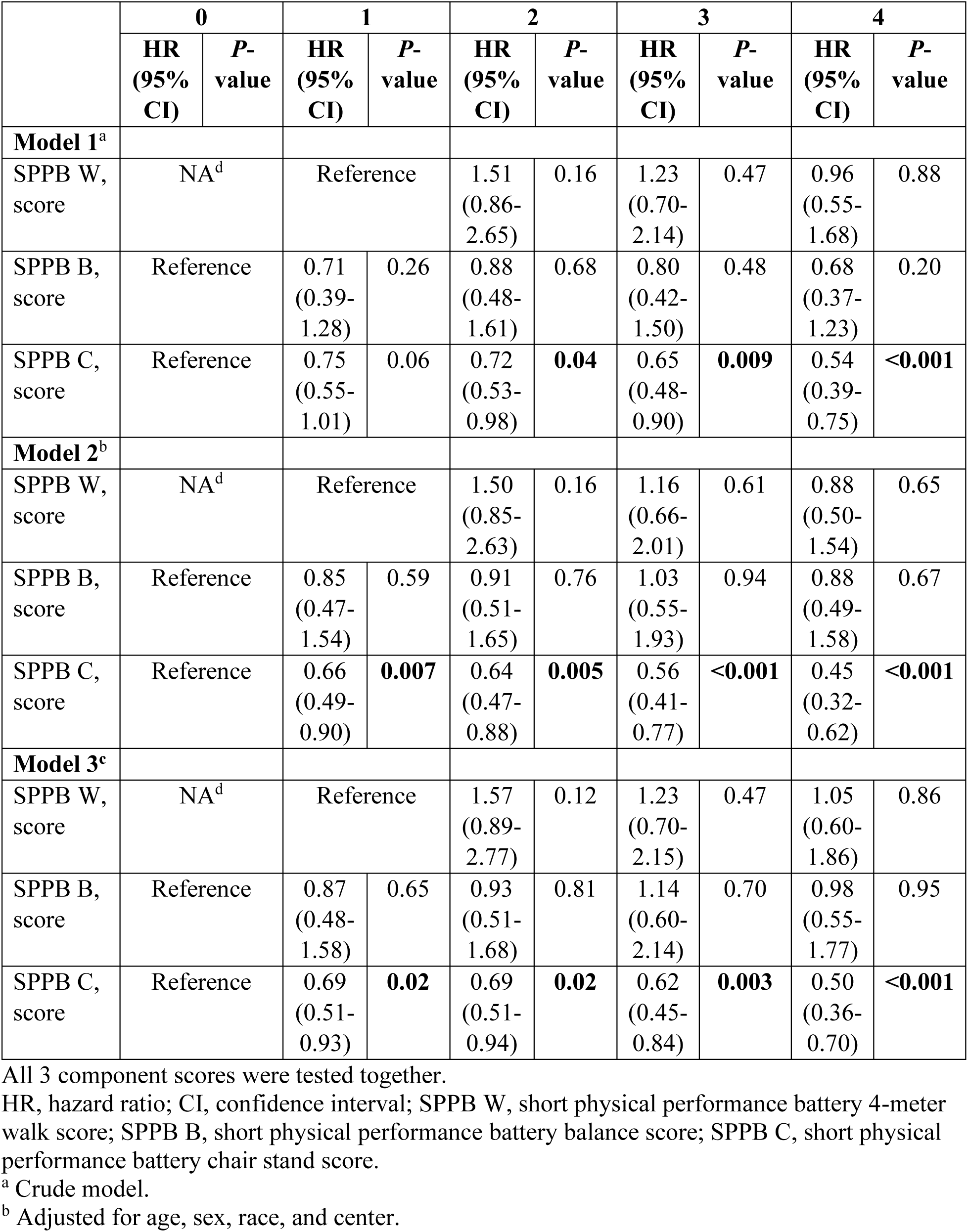
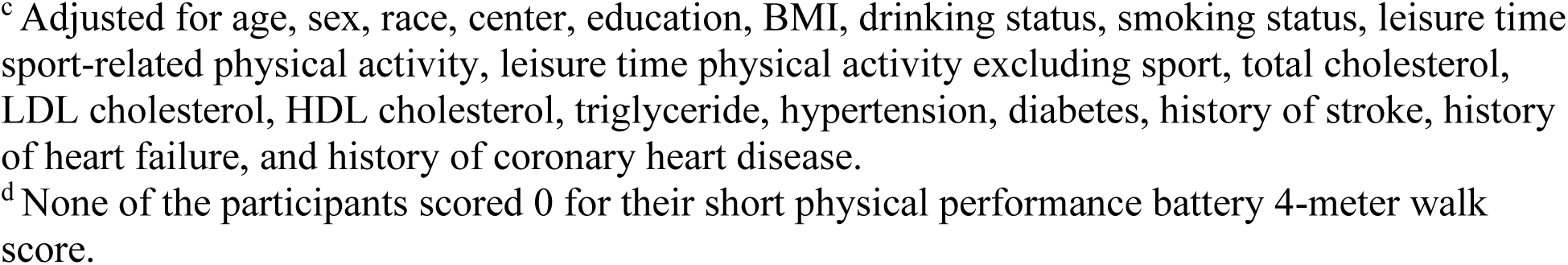
Association between Short Physical Performance Battery Component Scores and Risk of Incident Atrial Fibrillation.

|  | 0 |  | 1 |  | 2 |  | 3 |  | 4 |  |
| --- | --- | --- | --- | --- | --- | --- | --- | --- | --- | --- |
|  | HR<br>(95%<br>CI) | P-value | HR<br>(95%<br>CI) | P-value | HR<br>(95%<br>CI) | P-value | HR<br>(95%<br>CI) | P-value | HR<br>(95%<br>CI) | P-value |
| **Model 1<sup>a</sup>** |  |  |  |  |  |  |  |  |  |  |
| SPPB W,<br>score | NA<sup>d</sup> |  | Reference |  | 1.51<br>(0.86-2.65) | 0.16 | 1.23<br>(0.70-2.14) | 0.47 | 0.96<br>(0.55-1.68) | 0.88 |
| SPPB B,<br>score | Reference |  | 0.71<br>(0.39-1.28) | 0.26 | 0.88<br>(0.48-1.61) | 0.68 | 0.80<br>(0.42-1.50) | 0.48 | 0.68<br>(0.37-1.23) | 0.20 |
| SPPB C,<br>score | Reference |  | 0.75<br>(0.55-1.01) | 0.06 | 0.72<br>(0.53-0.98) | **0.04** | 0.65<br>(0.48-0.90) | **0.009** | 0.54<br>(0.39-0.75) | **<0.001** |
| **Model 2<sup>b</sup>** |  |  |  |  |  |  |  |  |  |  |
| SPPB W,<br>score | NA<sup>d</sup> |  | Reference |  | 1.50<br>(0.85-2.63) | 0.16 | 1.16<br>(0.66-2.01) | 0.61 | 0.88<br>(0.50-1.54) | 0.65 |
| SPPB B,<br>score | Reference |  | 0.85<br>(0.47-1.54) | 0.59 | 0.91<br>(0.51-1.65) | 0.76 | 1.03<br>(0.55-1.93) | 0.94 | 0.88<br>(0.49-1.58) | 0.67 |
| SPPB C,<br>score | Reference |  | 0.66<br>(0.49-0.90) | **0.007** | 0.64<br>(0.47-0.88) | **0.005** | 0.56<br>(0.41-0.77) | **<0.001** | 0.45<br>(0.32-0.62) | **<0.001** |
| **Model 3<sup>c</sup>** |  |  |  |  |  |  |  |  |  |  |
| SPPB W,<br>score | NA<sup>d</sup> |  | Reference |  | 1.57<br>(0.89-2.77) | 0.12 | 1.23<br>(0.70-2.15) | 0.47 | 1.05<br>(0.60-1.86) | 0.86 |
| SPPB B,<br>score | Reference |  | 0.87<br>(0.48-1.58) | 0.65 | 0.93<br>(0.51-1.68) | 0.81 | 1.14<br>(0.60-2.14) | 0.70 | 0.98<br>(0.55-1.77) | 0.95 |
| SPPB C,<br>score | Reference |  | 0.69<br>(0.51-0.93) | **0.02** | 0.69<br>(0.51-0.94) | **0.02** | 0.62<br>(0.45-0.84) | **0.003** | 0.50<br>(0.36-0.70) | **<0.001** |

All 3 component scores were tested together.
HR, hazard ratio; CI, confidence interval; SPPB W, short physical performance battery 4-meter walk score; SPPB B, short physical performance battery balance score; SPPB C, short physical performance battery chair stand score.
<sup>a</sup> Crude model.
<sup>b</sup> Adjusted for age, sex, race, and center.
<sup>c</sup> Adjusted for age, sex, race, center, education, BMI, drinking status, smoking status, leisure time sport-related physical activity, leisure time physical activity excluding sport, total cholesterol, LDL cholesterol, HDL cholesterol, triglyceride, hypertension, diabetes, history of stroke, history of heart failure, and history of coronary heart disease.
<sup>d</sup> None of the participants scored 0 for their short physical performance battery 4-meter walk score.

### Sex and Race Stratified Analyses

Among 2,824 female participants (416 incident AF events), higher SPPB was significantly associated with reduced risk of incident AF in the fully adjusted model (HR: 0.94, 95% CI: 0.90-0.98, p-value: 0.004), whereas other measures of physical function were not significantly associated with risk of incident AF (Table S1). Among 1,979 male participants (393 incident AF events), higher SPPB (HR: 0.91, 95% CI: 0.87-0.96, p-value: <0.001) and greater grip strength (HR: 0.85, 95% CI: 0.75-0.96, p-value: 0.009) were significantly associated with reduced risk of incident AF, while longer 4-meter walk time (HR: 1.18, 95% CI: 1.08-1.29, p-value: <0.001) was significantly associated with increased risk of incident AF, after fully adjusting for covariates (Table S1). The p-values for interaction between physical function and sex were not significant (SPPB and sex: 0.62; 4-meter walk time and sex: 0.07; grip strength and sex: 0.59) (Table S2).

Among 1,067 black participants (111 incident AF events), no significant associations between physical function and risk of incident AF were observed in the fully adjusted models (Table S3). In contrast, among 3,736 white participants (698 incident AF events), higher SPPB (HR: 0.92, 95% CI: 0.89-0.96, p-value: <0.001) and greater grip strength (HR: 0.88, 95% CI: 0.79-0.98, p-value: 0.02) were significantly associated with reduced risk of incident AF, while longer 4-meter walk time (HR: 1.09, 95% CI: 1.04-1.15, p-value: 0.001) was significantly associated with increased risk of incident AF, after fully adjusting for covariates (Table S3). However, the p-values for interaction between physical function and race were not significant (SPPB and race: 0.89; 4-meter walk time and race: 0.87; grip strength and race: 0.64) (Table S2).

### Nonlinear Association between Physical Function and Incident AF

The AIC values for determining the number of knots for the restricted cubic spline analysis are shown in Table S4. Based on these, 3 knots were used for SPPB and grip strength, whereas 4 knots for 4-meter walk time. Nonlinear associations between physical function and risk of incident AF are delineated in Figure 3. Specifically, 4-meter walk time had a significant nonlinear association with risk of incident AF in the fully adjusted model (p-value: 0.005) (Table S5). This significant nonlinear association was also observed in models 1 and 2. SPPB and grip strength showed significant overall associations with risk of incident AF (SPPB spline overall p-value: <0.001; grip strength spline overall p-value: 0.006), though no significant nonlinearity was detected (Table S5).

**Figure 3.**
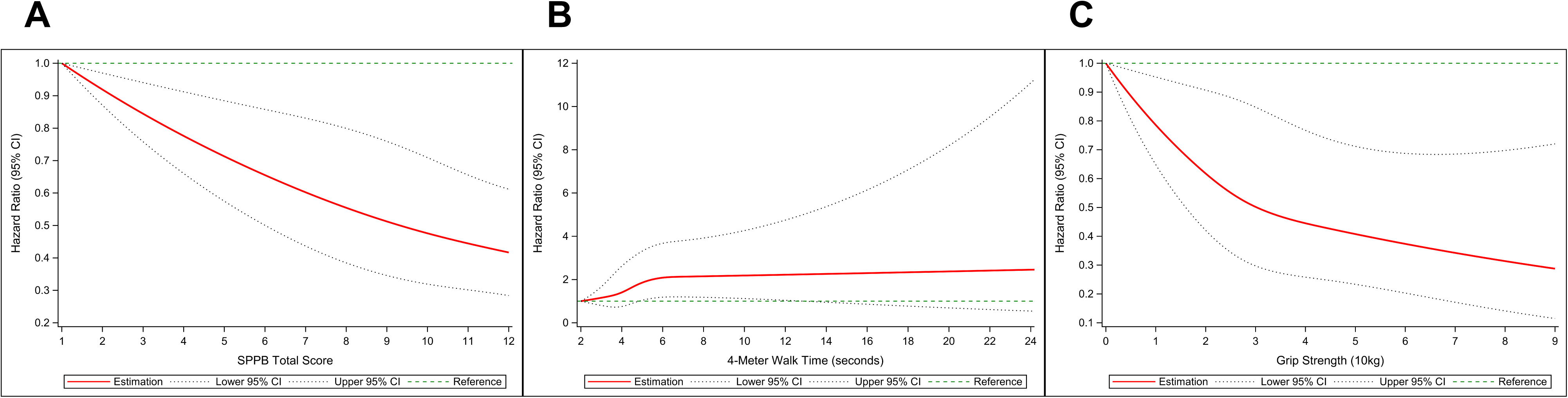
Fully adjusted dose-response associations between physical function and risk of incident atrial fibrillation using Cox regression models with restricted cubic splines: (A) SPPB; (B) 4-Meter Walk Time; (C) Grip Strength. For SPPB and Grip Strength, 3 knots placed at the 10^th^, 50^th^, and 90^th^ percentiles were used. For 4-Meter Walk Time, 4 knots placed at the 5^th^, 35^th^, 65^th^, and 95^th^ percentiles were used. CI, confidence interval; SPPB, short physical performance battery.

## Discussion

In this community-based cohort study investigating the association between physical function and risk of incident AF, all three measures of physical function (SPPB, 4-meter walk time, grip strength) were associated with risk of incident AF. Specifically, higher SPPB and greater grip strength were associated with reduced risk of incident AF, while longer 4-meter walk time was associated with increased risk of incident AF. These results are consistent with a previous study that found an inverse relationship between grip strength and incident AF risk.^22^ However, that study was limited by its small sample size and lack of incorporation of other physical function measures. The current study’s large sample size and consideration of multiple measures of physical function strengthen the evidence base. Among the physical function measures used in this study, higher SPPB showed the strongest association with reduced risk of incident AF. The current study uniquely highlights the importance of multi-domain measures of physical function such as SPPB. While our results demonstrate that better physical function is associated with reduced risk of incident AF, previous studies have found that AF is associated with reduced physical function,^35, 36^ suggesting a potential bidirectional relationship between physical function and AF.

This study has several clinical implications for the prevention of incident AF. Better physical function, as assessed by SPPB, 4-meter walk time, and grip strength, was independently associated with reduced risk of incident AF. Since physical function is modifiable, interventions focused on improving strength and mobility may represent a practical strategy for AF prevention. In our analysis, higher SPPB demonstrated the strongest association with reduced risk of incident AF, consistent with its multi-domain nature integrating balance, gait speed, and lower-extremity strength. In light of our findings, categorizing SPPB (low 0-6, intermediate 7-9, and high 10-12) may be more clinically relevant than treating it as a continuous variable. However, longer 4-meter walk time was independently associated with increased risk of incident AF, and greater grip strength was independently associated with reduced risk of incident AF. Thus, even in the absence of SPPB, 4-meter walk time and grip strength could provide valuable information in relation to risk of incident AF.

Several mechanisms could underlie the association between physical function and risk of incident AF. Measures of physical function may serve as indicators of the state of metabolic dysfunction, systemic inflammation, oxidative stress, and neurodegeneration, which could lead to AF.^37, 38, 39, 40, 41^ Particularly, muscle strength appears to be an important factor.^12, 15, 19, 22^ Muscle strength is a key component that is present in all three physical function measures used in this study. Furthermore, the SPPB component analysis showed that the chair stand component was the only SPPB component that consistently had a significant association with reduced risk of incident AF. Specifically, in the fully adjusted model, the maximum possible chair stand component score of 4 was associated with a 50% reduced risk of incident AF compared to the lowest possible score of 0. Given that the chair stand component assesses lower-body muscle strength, the current study suggests muscle strength could be an important driver of the association between physical function and risk of incident AF.

In analyses exploring nonlinearity, 4-meter walk time was the only measure of physical function that had a significant nonlinear association with risk of incident AF in multivariable models. Specifically, the hazard ratio increased at the lower range of 4-meter walk time but plateaued afterward. Such result implies that the discriminative value of 4-meter walk time stays within the lower range, which could explain the lack of statistical significance once SPPB was considered.

The current study has several strengths including the large sample size, extensive follow-up period, diverse study participants, multiple objectively measured physical function parameters, and incorporation of multiple covariates. Despite the notable strengths of this study, several limitations must be acknowledged. This study was limited in its ability to investigate potential mechanisms. Future mechanistic and randomized studies should attempt to investigate the direct pathways that explain the association between physical function and risk of incident AF. Furthermore, the lack of continuous electrocardiographic monitoring for identifying incident AF cases could lead to under-detection of subclinical and paroxysmal AF cases. Despite this limitation, prior research has demonstrated that hospital discharge codes and death certificates provide a reliable method for the ascertainment of incident AF.^26, 42^ Additionally, the study participants demonstrated good physical function, which may limit the generalizability of the findings to other, more frail populations. Although the associations between physical function and risk of incident AF were statistically significant, the effect sizes were small and should be interpreted with caution. Finally, the sample size was not sufficient for studying race-specific associations, and the possibility of residual confounding remains.

## Conclusion

The findings of this study have demonstrated that better physical function is associated with reduced risk of incident AF. Higher SPPB was most strongly associated with reduced risk of incident AF, though longer 4-meter walk time was independently associated with increased risk of incident AF, and greater grip strength was independently associated with reduced risk of incident AF. Such findings imply the potential of physical function as a novel modifiable target for AF prevention. Within SPPB, higher chair stand component score was significantly associated with reduced risk of incident AF, emphasizing the role of muscle strength in the association between physical function and risk of incident AF. Future studies should aim to investigate the mechanisms that explain the relationship between physical function and risk of incident AF, identify individuals at higher risk of incident AF based on levels of physical function, and develop targeted interventions for such high-risk individuals.

## Data Availability

Data are available from the Atherosclerosis Risk in Communities (ARIC) Study upon approval of a data use application; restrictions apply and data are not publicly available.

## Acknowledgements

The authors thank the staff and participants of the ARIC study for their important contributions.

## References

1. Kornej J, Börschel CS, Benjamin EJ, Schnabel RB. Epidemiology of Atrial Fibrillation in the 21st Century: Novel Methods and New Insights. Circ Res. 2020;127:4–20.

2. Linz D, Gawalko M, Betz K, Hendriks JM, Lip GYH, Vinter N, Guo Y, Johnsen S. Atrial fibrillation: epidemiology, screening and digital health. Lancet Reg Health Eur. 2024;37:100786.

3. Benjamin EJ, Muntner P, Alonso A, Bittencourt MS, Callaway CW, Carson AP, Chamberlain AM, Chang AR, Cheng S, Das SR, et al. Heart Disease and Stroke Statistics-2019 Update: A Report From the American Heart Association. Circulation. 2019;139:e56–e528.

4. Roth GA, Mensah GA, Johnson CO, Addolorato G, Ammirati E, Baddour LM, Barengo NC, Beaton AZ, Benjamin EJ, Benziger CP, et al. Global Burden of Cardiovascular Diseases and Risk Factors, 1990-2019: Update From the GBD 2019 Study. J Am Coll Cardiol. 2020;76:2982–3021.

5. Schnabel RB, Yin X, Gona P, Larson MG, Beiser AS, McManus DD, Newton-Cheh C, Lubitz SA, Magnani JW, Ellinor PT, et al. 50 year trends in atrial fibrillation prevalence, incidence, risk factors, and mortality in the Framingham Heart Study: a cohort study. Lancet. 2015;386:154–162.

6. Dilaveris PE, Kennedy HL. Silent atrial fibrillation: epidemiology, diagnosis, and clinical impact. Clin Cardiol. 2017;40:413–418.

7. Frederiksen TC, Dahm CC, Preis SR, Lin H, Trinquart L, Benjamin EJ, Kornej J. The bidirectional association between atrial fibrillation and myocardial infarction. Nat Rev Cardiol. 2023;20:631–644.

8. Wolf PA, Abbott RD, Kannel WB. Atrial fibrillation as an independent risk factor for stroke: the Framingham Study. Stroke. 1991;22:983–988.

9. Gopinathannair R, Chen LY, Chung MK, Cornwell WK, Furie KL, Lakkireddy DR, Marrouche NF, Natale A, Olshansky B, Joglar JA. Managing Atrial Fibrillation in Patients With Heart Failure and Reduced Ejection Fraction: A Scientific Statement From the American Heart Association. Circ Arrhythm Electrophysiol. 2021;14:HAE0000000000000078.

10. Rivard L, Friberg L, Conen D, Healey JS, Berge T, Boriani G, Brandes A, Calkins H, Camm AJ, Yee Chen L, et al. Atrial Fibrillation and Dementia: A Report From the AF-SCREEN International Collaboration. Circulation. 2022;145:392–409.

11. Hu X, Mok Y, Ding N, Sullivan KJ, Lutsey PL, Schrack JA, Palta P, Matsushita K. Physical Function and Subsequent Risk of Cardiovascular Events in Older Adults: The Atherosclerosis Risk in Communities Study. J Am Heart Assoc. 2022;11:e025780.

12. Leong DP, Teo KK, Rangarajan S, Lopez-Jaramillo P, Avezum A Jr, Orlandini A, Seron P, Ahmed SH, Rosengren A, Kelishadi R, et al. Prospective Urban Rural Epidemiology (PURE) Study investigators. Prognostic value of grip strength: findings from the Prospective Urban Rural Epidemiology (PURE) study. Lancet. 2015;386:266–273.

13. Ueno K, Kaneko H, Kamiya K, Itoh H, Okada A, Suzuki Y, Matsuoka S, Fujiu K, Michihata N, Jo T, et al. Clinical utility of simple subjective gait speed for the risk stratification of heart failure in a primary prevention setting. Sci Rep. 2022;12:11641.

14. Chainani V, Shaharyar S, Dave K, Choksi V, Ravindranathan S, Hanno R, Jamal O, Abdo A, Abi Rafeh N. Objective measures of the frailty syndrome (hand grip strength and gait speed) and cardiovascular mortality: a systematic review. Int J Cardiol. 2016;215:487–493.

15. Ding N, Ballew SH, Palta P, Schrack JA, Windham BG, Coresh J, Matsushita K. Muscle strength and incident cardiovascular outcomes in older adults. J Am Coll Cardiol. 2020;75:1090–1092.

16. Welsh CE, Celis-Morales CA, Ho FK, Brown R, Mackay DF, Lyall DM, Anderson JJ, Pell JP, Gill JMR, Sattar N, et al. Grip strength and walking pace and cardiovascular disease risk prediction in 406,834 UK Biobank participants. Mayo Clin Proc. 2020;95:879–888.

17. McGinn AP, Kaplan RC, Verghese J, Rosenbaum DM, Psaty BM, Baird AE, Lynch JK, Wolf PA, Kooperberg C, Larson JC, et al. Walking speed and risk of incident ischemic stroke among postmenopausal women. Stroke. 2008;39:1233–1239.

18. Imran TF, Orkaby A, Chen J, Selvaraj S, Driver JA, Gaziano JM, Djoussé L. Walking pace is inversely associated with risk of death and cardiovascular disease: the Physicians’ Health Study. Atherosclerosis. 2019;289:51–56.

19. Celis-Morales CA, Welsh P, Lyall DM, Steell L, Petermann F, Anderson J, Iliodromiti S, Sillars A, Graham N, Mackay DF, et al. Associations of grip strength with cardiovascular, respiratory, and cancer outcomes and all cause mortality: prospective cohort study of half a million UK Biobank participants. BMJ. 2018;361:k1651.

20. Zeki Al Hazzouri A, Mayeda ER, Elfassy T, Lee A, Odden MC, Thekkethala D, Wright CB, Glymour MM, Haan MN. Perceived walking speed, measured tandem walk, incident stroke, and mortality in older latino adults: a prospective cohort study. J Gerontol A Biol Sci Med Sci. 2017;72:676–682.

21. Quan M, Xun P, Wang R, He K, Chen P. Walking pace and the risk of stroke: a meta-analysis of prospective cohort studies. J Sport Health Sci. 2020;9:521–529.

22. Kunutsor SK, Mäkikallio TH, Jae SY, Khan H, Voutilainen A, Laukkanen JA. Handgrip Strength and Risk of Atrial Fibrillation. Am J Cardiol. 2020;137:135–138.

23. Wright JD, Folsom AR, Coresh J, Sharrett AR, Couper D, Wagenknecht LE, Mosley TH Jr, Ballantyne CM, Boerwinkle EA, Rosamond WD, et al. The ARIC (Atherosclerosis Risk In Communities) Study: JACC focus seminar 3/8. J Am Coll Cardiol. 2021;77:2939–2959.

24. Guralnik JM, Simonsick EM, Ferrucci L, Glynn RJ, Berkman LF, Blazer DG, Scherr PA, Wallace RB. A Short Physical Performance Battery assessing lower extremity function: association with self-reported disability and prediction of mortality and nursing home admission. J Gerontol. 1994;49:M85–M94.

25. Li D, Alam AB, Yu F, Kucharska-Newton A, Windham BG, Alonso A. Sphingolipids and physical function in the Atherosclerosis Risk in Communities (ARIC) study. Sci Rep. 2021;11:1169.

26. Alonso A, Agarwal SK, Soliman EZ, Ambrose M, Chamberlain AM, Prineas RJ, Folsom AR. Incidence of atrial fibrillation in whites and African-Americans: the Atherosclerosis Risk in Communities (ARIC) study. Am Heart J. 2009;158:111–117.

27. Baecke JA, Burema J, Frijters JE. A short questionnaire for the measurement of habitual physical activity in epidemiological studies. Am J Clin Nutr. 1982;36:936–942.

28. Rosamond WD, Folsom AR, Chambless LE, Wang CH, McGovern PG, Howard G, Copper LS, Shahar E. Stroke incidence and survival among middle-aged adults: 9-year follow-up of the Atherosclerosis Risk in Communities (ARIC) cohort. Stroke. 1999;30:736–743.

29. Loehr LR, Rosamond WD, Chang PP, Folsom AR, Chambless LE. Heart failure incidence and survival (from the Atherosclerosis Risk in Communities study). Am J Cardiol. 2008;101:1016–1022.

30. White AD, Folsom AR, Chambless LE, Sharret AR, Yang K, Conwill D, Higgins M, Williams OD, Tyroler HA. Community surveillance of coronary heart disease in the Atherosclerosis Risk in Communities (ARIC) Study: methods and initial two years’ experience. J Clin Epidemiol. 1996;49:223–233.

31. Guralnik JM, Ferrucci L, Simonsick EM, Salive ME, Wallace RB. Lower-extremity function in persons over the age of 70 years as a predictor of subsequent disability. N Engl J Med. 1995;332:556–561.

32. Westerman S, Wenger N. Gender Differences in Atrial Fibrillation: A Review of Epidemiology, Management, and Outcomes. Curr Cardiol Rev. 2019;15:136–144.

33. Tamirisa KP, Al-Khatib SM, Mohanty S, Han JK, Natale A, Gupta D, Russo AM, Al-Ahmad A, Gillis AM, Thomas KL. Racial and Ethnic Differences in the Management of Atrial Fibrillation. CJC Open. 2021;3:S137–S148.

34. Desquilbet L, Mariotti F. Dose-response analyses using restricted cubic spline functions in public health research. Stat Med. 2010;29:1037–1057.

35. Ceolin C, Mizzon E, Noale M, Ravelli A, Pigozzo S, Curreri C, Zanforlini BM, Manzato E, Coin A, Devita M, et al. The Impact of Atrial Fibrillation on Physical Performance in Older Adults: A Longitudinal Study in Relation to Cognitive Function. J Am Med Dir Assoc. 2025;26:105764.

36. Magnani JW, Wang N, Benjamin EJ, Garcia ME, Bauer DC, Butler J, Ellinor PT, Kritchevsky S, Marcus GM, Newman A, et al. Atrial Fibrillation and Declining Physical Performance in Older Adults: The Health, Aging, and Body Composition Study. Circ Arrhythm Electrophysiol. 2016;9:e003525.

37. Forman DE, Arena R, Boxer R, Dolansky MA, Eng JJ, Fleg JL, Haykowsky M, Jahangir A, Kaminsky LA, Kitzman DW, et al. Prioritizing functional capacity as a principal end point for therapies oriented to older adults with cardiovascular disease: a scientific statement for healthcare professionals from the American Heart Association. Circulation. 2017;135:e894–e918.

38. López-Otín C, Blasco MA, Partridge L, Serrano M, Kroemer G. The hallmarks of aging. Cell. 2013;153:1194–1217.

39. Van Wagoner DR. Oxidative stress and inflammation in atrial fibrillation: role in pathogenesis and potential as a therapeutic target. J Cardiovasc Pharmacol. 2008;52:306–313.

40. Chu H, Guo X, Xu HC, Wang S, Guo Z, Tian Y, Wang Y. A causal relationship between functional connectivity of brain networks and cardiovascular disease: A Mendelian randomization study. Medicine (Baltimore). 2025;104:e43131.

41. Bode D, Pronto JRD, Schiattarella GG, Voigt N. Metabolic remodelling in atrial fibrillation: manifestations, mechanisms and clinical implications. Nat Rev Cardiol. 2024;21:682–700.

42. Jensen PN, Johnson K, Floyd J, Heckbert SR, Carnahan R, Dublin S. A systematic review of validated methods for identifying atrial fibrillation using administrative data. Pharmacoepidemiol Drug Saf. 2012;21:141–147.

