## Supplemental Material for "Association between Physical Function and Incidence of Atrial Fibrillation: The Atherosclerosis Risk in Communities (ARIC) Study"

**Table S1**. Sex Differences in Association between Measures of Physical Function and Risk of Incident Atrial Fibrillation

|  | **Model 1**^a^ | | | **Model 2**^b^ | | | **Model 3^c^** | | |
| --- | --- | --- | --- | --- | --- | --- | --- | --- | --- |
|  | **HR** | **95% CI** | ***P*-value** | **HR** | **95% CI** | ***P*-value** | **HR** | **95% CI** | ***P*-value** |
| **Females (n=2824; 416 AF events)** |  |  |  |  |  |  |  |  |  |
| SPPB, score | 0.89 | 0.86-0.92 | **<0.001** | 0.90 | 0.86-0.93 | **<0.001** | 0.94 | 0.90-0.98 | **0.004** |
| 4-meter walk time, seconds | 1.12 | 1.07-1.17 | **<0.001** | 1.10 | 1.05-1.16 | **<0.001** | 1.05 | 0.99-1.11 | 0.14 |
| Grip strength, 10 kg | 0.71 | 0.61-0.83 | **<0.001** | 0.86 | 0.71-1.03 | 0.09 | 0.90 | 0.75-1.08 | 0.24 |
| **Males (n=1979; 393 AF events)** |  |  |  |  |  |  |  |  |  |
| SPPB, score | 0.87 | 0.83-0.91 | **<0.001** | 0.88 | 0.84-0.92 | **<0.001** | 0.91 | 0.87-0.96 | **<0.001** |
| 4-meter walk time, seconds | 1.21 | 1.14-1.29 | **<0.001** | 1.23 | 1.14-1.33 | **<0.001** | 1.18 | 1.08-1.29 | **<0.001** |
| Grip strength, 10 kg | 0.69 | 0.62-0.78 | **<0.001** | 0.79 | 0.70-0.89 | **<0.001** | 0.85 | 0.75-0.96 | **0.009** |

Each measure of physical function was tested separately.

HR, hazard ratio; CI, confidence interval; SPPB, short physical performance battery (total score).

^a^ Crude model.

^b^ Adjusted for age, race, and center.

^c^ Adjusted for age, race, center, education, BMI, drinking status, smoking status, leisure time sport-related physical activity, leisure time physical activity excluding sport, total cholesterol, LDL cholesterol, HDL cholesterol, triglyceride, hypertension, diabetes, history of stroke, history of heart failure, and history of coronary heart disease.

**Table S2**. P-values for Interactions between Measures of Physical Function and Sex or Race on Risk of Incident Atrial Fibrillation

|  | ***P*-value for Interaction with Sex^a^** | ***P*-value for Interaction with Race^a^** |
| --- | --- | --- |
| SPPB, score | 0.62 | 0.89 |
| 4-meter walk time, seconds | 0.07 | 0.87 |
| Grip strength, 10 kg | 0.59 | 0.64 |

Each measure of physical function was tested separately.

SPPB, short physical performance battery (total score).

^a^ Model included the main effects, interaction term, and were adjusted for age, center, education, BMI, drinking status, smoking status, leisure time sport-related physical activity, leisure time physical activity excluding sport, total cholesterol, LDL cholesterol, HDL cholesterol, triglyceride, hypertension, diabetes, history of stroke, history of heart failure, and history of coronary heart disease. When assessing interaction by sex, race was adjusted; when assessing interaction by race, sex was adjusted.

**Table S3**. Racial Differences in Association between Measures of Physical Function and Risk of Incident Atrial Fibrillation

|  | **Model 1**^a^ | | | **Model 2**^b^ | | | **Model 3^c^** | | |
| --- | --- | --- | --- | --- | --- | --- | --- | --- | --- |
|  | **HR** | **95% CI** | ***P*-value** | **HR** | **95% CI** | ***P*-value** | **HR** | **95% CI** | ***P*-value** |
| **Black (n=1067; 111 AF events)** |  |  |  |  |  |  |  |  |  |
| SPPB, score | 0.87 | 0.81-0.92 | **<0.001** | 0.88 | 0.82-0.94 | **<0.001** | 0.96 | 0.89-1.04 | 0.35 |
| 4-meter walk time, seconds | 1.14 | 1.06-1.23 | **<0.001** | 1.13 | 1.04-1.22 | **0.004** | 1.02 | 0.91-1.13 | 0.76 |
| Grip strength, 10 kg | 0.83 | 0.68-1.02 | 0.07 | 0.76 | 0.59-0.97 | **0.03** | 0.84 | 0.66-1.08 | 0.17 |
| **White (n=3736; 698 AF events)** |  |  |  |  |  |  |  |  |  |
| SPPB, score | 0.87 | 0.84-0.89 | **<0.001** | 0.89 | 0.86-0.92 | **<0.001** | 0.92 | 0.89-0.96 | **<0.001** |
| 4-meter walk time, seconds | 1.15 | 1.11-1.19 | **<0.001** | 1.13 | 1.08-1.18 | **<0.001** | 1.09 | 1.04-1.15 | **0.001** |
| Grip strength, 10 kg | 0.98 | 0.91-1.05 | 0.50 | 0.82 | 0.74-0.92 | **<0.001** | 0.88 | 0.79-0.98 | **0.02** |

Each measure of physical function was tested separately.

HR, hazard ratio; CI, confidence interval; SPPB, short physical performance battery (total score).

^a^ Crude model.

^b^ Adjusted for age, sex, and center.

^c^ Adjusted for age, sex, center, education, BMI, drinking status, smoking status, leisure time sport-related physical activity, leisure time physical activity excluding sport, total cholesterol, LDL cholesterol, HDL cholesterol, triglyceride, hypertension, diabetes, history of stroke, history of heart failure, and history of coronary heart disease.

**Table S4**. Comparison of Akaike Information Criterion with Different Number of Knots

|  | **3 knots** | **4 knots** | **5 knots** |
| --- | --- | --- | --- |
|  | **AIC^a^** | **AIC^a^** | **AIC^a^** |
| SPPB, score | **13003.700** | 13005.533 | 13006.925 |
| 4-meter walk time, seconds | 13004.468 | **13003.491** | 13004.965 |
| Grip strength, 10 kg | **13013.915** | 13014.645 | 13015.704 |

Each measure of physical function was tested separately.

AIC, akaike information criterion; SPPB, short physical performance battery (total score).

^a^ Adjusted for age, sex, race, center, education, BMI, drinking status, smoking status, leisure time sport-related physical activity, leisure time physical activity excluding sport, total cholesterol, LDL cholesterol, HDL cholesterol, triglyceride, hypertension, diabetes, history of stroke, history of heart failure, and history of coronary heart disease.

**Table S5**. Nonlinear Association between Measures of Physical Function and Risk of Incident Atrial Fibrillation Using Restricted Cubic Splines

|  | **Model 1**^a^ | | **Model 2**^b^ | | **Model 3^c^** | |
| --- | --- | --- | --- | --- | --- | --- |
|  | **Overall *P*-value** | **Nonlinear *P*-value** | **Overall *P*-value** | **Nonlinear *P*-value** | **Overall *P*-value** | **Nonlinear *P*-value** |
| SPPB, score | **<0.001** | 0.96 | **<0.001** | 0.97 | **<0.001** | 0.76 |
| 4-meter walk time, seconds | **<0.001** | **<0.001** | **<0.001** | **<0.001** | **<0.001** | **0.005** |
| Grip strength, 10 kg | 0.08 | 0.39 | **<0.001** | 0.43 | **0.006** | 0.29 |

Each measure of physical function was tested separately.

SPPB, short physical performance battery (total score).

^a^ Crude model.

^b^ Adjusted for age, sex, race, and center.

^c^ Adjusted for age, sex, race, center, education, BMI, drinking status, smoking status, leisure time sport-related physical activity, leisure time physical activity excluding sport, total cholesterol, LDL cholesterol, HDL cholesterol, triglyceride, hypertension, diabetes, history of stroke, history of heart failure, and history of coronary heart disease.
